# Service availability, readiness and health-seeking behavior of gender based violence survivor services in primary healthcare facilities in Dodoma, Tanzania. An Explanatory-sequential study design

**DOI:** 10.1101/2024.06.09.24308665

**Authors:** Johnson W. Mwamasangula, Nyasiro S. Gibore

**Affiliations:** Department of Public Health and Community Nursing, University of Dodoma

**Keywords:** gender-based violence survivor’s services, health-seeking behavior, service availability and readiness, primary healthcare

## Abstract

**Background:** The health sector is a key stakeholder in GBV national response network however, delivery of quality healthcare services for GBV survivors is highly neglected.

**Methods:** This study will use a mixed-research approach by employing an explanatory sequential study design. Phase one will involve a cross-sectional survey of 61 primary healthcare facilities to examine availability and readiness of GBV survivor’s services. The second phase will involve a descriptive qualitative study among healthcare providers, community healthcare workers and clients to examine the health-seeking behavior and challenges in providing and receiving GBV survivor’s services in primary healthcare facilities. The study will be conducted between March-April 2024 in Dodoma City Council, Tanzania.

**Discussion:** This study will have potential implications to program managers, policy makers and healthcare providers to tailor interventions that will improve the quality of healthcare service delivery especially towards integrated GBV survivor’s services. But also, understanding the health-seeking behavior and challenges associated with access and provision of GBV survivor’s services will help unveiling gaps in the clients’ point of view and the community, their needs for the GBV survivor’s services and enhancing client-healthcare provider relationship.

## Background

Gender-based violence (GBV) comprises acts of cruelty and maltreatment directed against a person due to their sex or gender status by inflicting harm (sexual, emotional, physical), threats and deprivations of liberty (1,2). GBV doesn’t have boundaries thus it affects all people but girls and women are more victims of GBV forms compares to men (3,4). Gender-based violence (GBV), which is an expression of gender inequality and toxic masculinity (González & Rodríguez-Planas, 2020) is a public health problem that is present in all societies at different degrees of prevalence and severity (1) but occurrences of gender-based violence are highly reported in low-middle income countries and in those countries with high prevalence of human immunodeficiency virus (HIV) (5).

Several manifestations of gender based violence exist such as intimate partner violence (IPV), domestic violence, femicides and honor killings, female genital mutilation (FGM), child early and forced marriage and online or technology facilitated violence (6). The acts of gender based violence can be perpetrated as physical, sexual, psychological or economically by manifesting control over a person’s mobility and agency (6–9). Countries are responsible to ensure the fulfillment of SDG 5 by ensuring that communities especially vulnerable groups like women have access to inclusive comprehensive GBV prevention and response services in healthcare facilities by ensuring the operationalization of the four basic referral pathways for all survivors which are psychosocial, legal, safety and medical (2,10).

Globally, approximately 736 million women equivalent to one in three or 30% aged 15 and older are exposed to various forms of gender based violence especially physical and/or sexual intimate partner violence, non-partner sexual violence, or both at least once in their lifetime (8,9,11–13). Roughly, the prevalence of GBV among men is 1 in 9 men with unwanted sexual contact ranking in their lives (14). GBV contributes to adverse impacts in economic development of many countries that have weak protective laws against gender-based violence, countries with wealth stemming from natural resource, in countries where decision making potential is limited and during economic and natural disaster crisis such as COVID-19 (15).

South Asia and Sub-Saharan Africa are the regions with high prevalence of GBV whereby it is estimated that more than 30% of women of reproductive age experience gender based violence in their lifetime such as IPV (12). Among teenagers, IPV is a most common form of GBV whereby 1 in 5 of teenagers are experiencing IPV with those aged 15 to 19 being three times more likely than women over 45 to face IPV (3,12,16,17). The prevalence of gender-based violence in sub-Saharan Africa ranges from 11.6% to 75.6% due to paucity and sensitivity of the issue in the region as it is deeply rooted in social structures (3,5). Socio-demographic factors, individual factors, family factors, spouse’s/partner’s habit, and experience in the past plays a key role in the existence of GBV in the region (3). In East Africa, women’s experiences of sexual violence are twice than men, whereby physical violence is more pervasive than sexual violence at 50% in Burundi, 44% in Tanzania, 35% in Rwanda and 14% in Kenya of women aged 15-49 (18).

In Tanzania, GBV prevalence is pervasive whereby 40% of all women of reproductive age have experienced physical violence and 17% have experienced sexual violence (19–21). In realization of the significant detrimental impacts of GBV in Tanzania, the government has increasingly adopting national policies and programs to integrate GBV services into existing public sector health services (22). According to reports by CAG, LHRC, TDHS 2022 and One Plan III, most acts of GBV are physical and sexual violence (rape and sodomy) with leading regions being Arusha, Dar es Salaam, Dodoma, Tanga, Katavi, Shinyanga, Mwanza, Morogoro, Mara, Kagera and Mbeya (19,21,23,24). Majority of GBV perpetrators are husbands/intimate partners (81%), relatives (26%), family friends (23%), strangers (22%) former husband/intimate partner (18%) (19). Power is highly associated with GBV acts in Tanzania as perpetrators are often bearing real or perceived power, decision making, or authority which they use to exercise control over their survivors (25) which subjects them into difficulty in getting help (26).

The global community and countries are committed into ensuring fulfillment of international, regional and national agreements to achieve gender equality and ending violence such as 2030 Agenda SDG 3 & 5 (10) and the National Action Plan to End Violence Against Women and Children (27). However, despite realization of the GBV as a public health problem, there is a gap in the quality of healthcare services for GBV survivors surrounded with variation in service delivery and lacking of data on GBV services in healthcare settings in the country (5). Most of GBV survivors seek health care services at primary level health facilities, but they experience challenges such as not receiving proper GBV services, delay in accessing or incomplete health care services (28). But also, other challenges are faced by healthcare workers and are not paid attention such as issues related to disclosure, exemption and policy gaps which affects the quality of GBV survivor’s services and health-seeking behavior (29).This leads to a question whether the hospital GBV services and programs meet clients’ needs and if they are available and ready (30).

Gaining familiarity about GBV service delivery is vital for program managers, policy makers and healthcare providers to tailor interventions that will improve the quality of healthcare service delivery and play part in combating various outcomes of GBV such as HIV/AIDS (5,18). But also, understanding the health-seeking behavior and challenges associated with access and provision of GBV survivor’s services is crucial in understanding the gap in the clients’ point of view and the community, their needs for the GBV survivor’s services and enhancing client-healthcare provider relationship.

## Methods/Design

### Study aim

The purpose of this study is to examine service availability and readiness of primary healthcare facilities to provide GBV survivor’s services. In addition, this study will examine the health-seeking behavior and challenges in providing gender based violence survivor’s services among primary healthcare facilities.

### Study design

A descriptive cross-sectional design and a descriptive qualitative design by using a mixed-research approach will be used.

### Study area

The study will be conducted in Dodoma City Council in Tanzania. Dodoma City Council has one parliament constituency and four divisions (4); *Dodoma Mjini, Hombolo, Kikombo* and *Zuzu* which hosts forty-one (41) wards. The council is in contract with St. Gemma Hospital which is used as a District hospital, 4 Health Centers and 32 Dispensaries. According to reports by CAG, LHRC, TDHS 2022 and One Plan III, most acts of GBV are physical and sexual violence (rape and sodomy) with Dodoma being among of the leading regions in the country thus requires an immediate action in ensuring services for GBV survivors are initiated from the primary level of healthcare delivery system (19,21,23,24).

### Study population and recruitment

This study will involve five types of study populations: all healthcare facilities at the primary level in the quantitative design and on qualitative design it will be healthcare providers working directly in GBV services, community healthcare workers, village officers, clients visiting the healthcare facility on the time of the study and cases or survivors who had previously received services in the selected primary healthcare facilities. Recruitment of study participants will start June 15-June 20, 2024. Healthcare provider who works directly on GBV services (doctor, nurse and social welfare officer) who will be available at the time of data collection or who will make an appointment will be recruited in the study. Community healthcare workers and village leaders will be contacted by telephone and letters to participate voluntarily in the study whereby those who will consent and be available at the time of data collection will be included in the study. All clients who will attend at the selected primary healthcare facility for healthcare services will be approached to participate in the study whereby those who will consent will be included in the study. Survivors of gender-based violence will be contacted through healthcare workers and asked to participate whereby those who will consent will be included in the study. The healthcare workers, village leaders, community healthcare workers and eligible study participants who will be seriously sick, mental illness problems and those enrolled in the similar study will be excluded in this study. All study participants will be communicated and recruited upon the arrival in the study area.

### Sampling and Sample size estimation

#### Sample size estimation of healthcare facilities

The WHO formula for sampling healthcare facilities will be used. According to the formula, when assessing lower health facility levels that are100 or fewer, investigator (s) should study all of them and when facilities are more than 100 investigator (s) should take at least 30% (WHO, 2009; Seif & Rashid, 2023). Total of 61 primary healthcare facilities (private and public) facilities found in the Dodoma City council will be included in the study.

On the descriptive qualitative phase, the sample size will depend on the saturation level of the collected information whereby a convenience sampling method will be used to select healthcare workers and a purposive sampling method will be used to select clients and community healthcare workers.

### Measurement of Variables

**GBV survivor’s service availability** refers to the comprehensiveness to which specific services for GBV survivors are available in a healthcare setting whenever and wherever they are required. This variable will be measured by an observational index using the three areas of tracer indicators which are infrastructure, workforce/core health personnel and services. This will be through expressing the indicators as a percentage score and then taking the mean of the area scores. Measurement will be by 11 items on a binary scale on the availability of well-functioning services, workforce and infrastructure. Each domain will comprise 3.6 marks whereby, the services domain will have 6 items, infrastructure domain 4 items and workforce 1 item. The total score will be 100, whereby each item will comprise of 9.09 marks. The score of 50 and above will indicate availability of GBV survivor’s services while the score below 50 will indicate unavailability of GBV survivor’s services. This cut-off point of 50% was also used in previous studies (31,32).

**GBV survivor’s service readiness** is the ability of the facility to respond to GBV survivors in a survivor-centred approach by having the necessary guidelines and utilities to facilitate service provision e.g. diagnostics, staff, guidelines, medicines, equipment and commodities to deliver GBV survivor’s service. This variable will be measured by observation of five domains/signal functions which are guidelines (standard precautions), trained staff, basic equipment, diagnostics (laboratory tests) and medicines/commodities. GBV survivor’s service readiness will be described by an index using the five general service readiness domains according to SARA Ver. 2.2 (33). A score will be generated per domain based on the number of domain elements present, then an overall general readiness score will be calculated based on the mean of the five domains. Measurement will be by 21 items on a binary scale. Each domain will have 4.2 marks whereby, guidelines domain will comprise 8 items, 6 items in laboratory and diagnostics domain, 4 items in training domain, 2 in equipments domain and 1 in management domain. Criteria to decide on the service readiness will be based on the total marks of 100 whereby each item will comprise 4.7 marks. The score of 50 will indicate readiness while the score below 50 will indicate not ready status of the primary healthcare facility to provide GBV survivor’s services. The cut-off point is based on the previous studies (31,32).

### Data collection procedure

Data collection will be conducted in the selected primary healthcare facilities in a 1-month timeframe by the Principal investigator assisted with two research assistants. Before engaging in the process of data collection, the research assistants will be trained on how to conduct the study and the ethical principles to adhere to during the whole time of the study. All information required for this study will be gathered on the arrival at the study area. In this study, several methods of data collection will be used depending on the type of data needed to answer a particular study objective as follows;

#### Survey

The survey method will be used to collect information on social demographic characteristics of study participants and information on availability and readiness of primary healthcare facilities to provide gender-based violence services

#### Documentary review

This method of data collection will be used to collect information on the management of GBV in lieu to Tanzania standard treatment guidelines in primary healthcare facilities as well as to review the information on the availability and readiness of primary healthcare facilities to provide gender-based survivor services.

#### Observation

This method will be used to collect information from registers, forms, available services and equipments, medicines and existing guidelines. Through an observation checklist, this method will collect data on the objective one; availability of GBV survivor’s services and objective two; readiness of healthcare facilities to provide GBV survivor’s services.

#### In-depth interview

This method will be used to record interviewees’ opinions and understanding on the health-seeking behavior and challenges faced by healthcare facilities to provide GBV survivor’s services. Due to its adaptability, the researcher will modify or delete certain questions to obtain the information required. This method will be used to the key informants on the provision of GBV survivor’s services on the objective three; health-seeking behavior for GBV survivor’s services and objective four; challenges in provision of GBV survivor’s services.

#### Focus Group Discussion

In this study, 10 FGDs will be conducted with the clients (5 for men, 5 for women) comprising of seven to nine respondents. In all discussions, tape recorder and note books will be used to record primary data given by different clients/members. Participants in the focus group discussions will be approached face to face through the guidance of a healthcare worker or a community healthcare workers (CHWs). This method will be used to collect data on the objective three and four among clients and community healthcare workers.

### Tools

This study will use a Questionnaire and an Observational checklist to collect data. The tools will be adapted from WHO’s HHFA 2023 (34) and the National Management Guidelines for the Health Sector Response to and Prevention of Gender-Based Violence (GBV) (25).

### Data management plan

Data obtained from participants will be stored in a secured computer with password that only the researcher’s team will be able to access, and all University of Dodoma safety protocols will be followed in order to secure participants information.

### Data analysis plan

For a **cross-sectional study design**, a Statistical Package for the Social Sciences (SPSS) ver. 24 will be used for data analysis. The designated level of the primary healthcare facility will be considered when performing the analyses as the findings will also be presented independently in respect of the level of the healthcare facility. Descriptive statistics will be used to analyze data collected on the level of availability and readiness of healthcare facilities to provide GBV survivor’s services through calculating the mean and present them in percentages and frequencies.

For a **descriptive qualitative design**, Interview and FGD data will be transcribed verbatim and translated into English language by two research assistants who are fluent in both Kiswahili and English. Thematic analysis will be done with the principal investigator who with the assistance of research assistance will proofread the transcripts to check for accuracy and import them into NVivo. 12 software for the coding process. The analysis will be carried out by reading the transferred transcripts and creating nodes in the process to get relevant texts from the transcripts. In the second step, codes will be assembled to form different themes whereby a thematic chart will be developed. The principal investigator will also check the accuracy of themes descriptions to find out if they had not been under or over represented. The third step will involve contextualizing the original codes along with the text so as to answer the aim of the study. Descriptive text will be used on the analytical themes by quotes which will illustrate the text in order to convey its meaning. Finally, results from FGDs and IDIs will be triangulated to identify the main themes.

### Techniques to enhance trustworthiness

Credibility will be ensured by triangulation of data collection methods (FGDs and IDIs), tools (voice recorder and field notes) and two research assistants. To ensure this principle is fulfilled, also the researcher will make attentive efforts to dispute any possible common assumptions that come at face value. Transferability will be ensured by thick descriptions of the research methodology by providing comprehensive information about the study, employing purposeful sampling and encouragement of confidentiality and privacy during the study. Dependability will be ensured by incorporating new issues that are emerging during the study by using an emerging design analysis. Also, more than one researcher in the study process will be used to ensure that the interpretations emerged in data through researchers’ triangulation and to enhance dependability of the data sources. Furthermore, the principal investigator is a Kiswahili native speaker and an expert in sociology whereby he will also be involved in data collection. Confirmability will be ensured through member checking technique during data collection and summarization or paraphrasing of the information received from IDIs and FGDs to ensure that what is heard or written down from the interviews and discussion is accurate.

### Ethical consideration and declaration

Ethical clearance was obtained from The University of Dodoma Research and Ethical Clearance Committee with Ref No. MA. 84/261/75/36, dated 08, May 2024. Ethical permission for this study was obtained from the University of Dodoma Research Ethics Committee (UDOM-REC). A written and verbal consent will be given to all study participants (health care providers, community healthcare workers, village executive officers, gender-based violence survivors and clients after explaining the purpose of the study and being told that their participation is voluntary and they can withdrawal from the study at any time.

### Dissemination plan

Findings obtained from this study will be disseminated to the University of Dodoma, the Ministry of Health, PO-RALG and will be submitted for publication in peer-reviewed journal.

### How amendments to the study, including termination, will be dealt with

If any changes are made to the study, including the study’s termination, this will be communicated to the journal auditoria, which will submit the changes or provide a reason for the termination.

## Data Availability

No datasets were generated or analysed during the current study. All relevant data from this study will be made available upon study completion.

## Supporting information

**S1 File**. This is an observational checklist for the availability and readiness of primary healthcare facilities to provide GBV survivor’s services

**S2 File**. This is a key informants in-depth interview guide combined with FGD guide for clients and community healthcare workers for examining health-seeking behavior and challenges in access and provision of GBV survivor’s services in primary healthcare facilities.

## List of abbreviations

CAG: Controller and Auditor General
CHWs: Community healthcare workers
FGD: Focus Group Discussion
FGM: Female Genital Mutilation
GBV: Gender-based violence
HCWs: Healthcare workers
HHFA: Harmonized Health Facility Assessment
HIV/AIDS: Human Immunodeficiency Virus/Acquired Immunodeficiency Syndrome
IDI: In-Depth Interview
IPV: Intimate Partner Violence
LHRC: Legal and Human Rights Center
MoH: Ministry of Health
MoHCDGEC: Ministry of Health, Community Development, Gender, Elderly and Children
PF 3: Police Form 3
SDGs: Sustainable Development Goals
SPSS: Statistical Package for the Social Sciences
TDHS: Tanzania Demographic Health Survey
UDOM-REC: University of Dodoma Research Ethics Committee
UNHCR: United Nations High Commission for Refugees
UNICEF: United Nations Children’s Fund
URT: United Republic of Tanzania
WHO: World Health Organization

## Consent for publication

Not applicable.

## Availability of data and materials

The datasets used and/or analyzed during the current study are available from the corresponding author on a reasonable request.

## Authors contribution

Writing – original draft, Conceptualization and Methodology: JWM

Writing – review, editing and Supervision: NSG

